# Comparison of Long-term Outcomes of Early Surgery Versus Conventional Treatment for Asymptomatic Severe Mitral Regurgitation: A Propensity Analysis

**DOI:** 10.1101/2025.03.11.25323798

**Authors:** Sung-Ji Park, Mijin Kim, Jihee Son, Ha Hye Jo, Ga Yun Kim, Jihoon Kim, Byung Joo Sun, Eun-Kyung Kim, Sahmin Lee, Jae Suk Yoo, Sung-Cheol Yun, Sung-Ho Jung, Jong-Min Song, Duk-Hyun Kang

## Abstract

**Background:** The timing of surgery in asymptomatic severe mitral regurgitation (MR) remains controversial. This study sought to compare long-term outcomes of early surgery with a conventional treatment strategy in asymptomatic patients with severe MR.

**Methods:** From 1996 to 2016, a total of 1,063 consecutive asymptomatic patients (673 men, age; 51±14 years) with severe degenerative MR and preserved left ventricular function were enrolled, and followed prospectively for a median of 12 years (interquartile range: 8 to 17 years). Early surgery was performed on 545 patients, while the conventional treatment strategy was chosen for 518. We compared overall and cardiac mortality between these two treatment strategies using propensity score adjustment.

**Results:** In the early surgery group, there was no operative mortality and mitral valve repair was successfully performed in 97% of patients. During follow-up, 8 patients (1.5%) in the early surgery group and 54 (10.4%) in the conventional management group died from cardiovascular causes (hazard ratio [HR], 0.17; 95% CI, 0.15 to 0.64; P = 0.001). A total of 74 deaths (13.6%) from any cause occurred in the early surgery group, whereas 116 (22.4%) occurred in the conventional management group (HR, 0.72; 95% CI, 0.52 to 0.99; P = 0.046). For the 358 propensity-score matched pairs, the early surgery group had a significantly lower risk of cardiac mortality than the conventional treatment group (HR, 0.18; 95% CI, 0.08 to 0.43; P < 0.001) and significantly lower cardiac mortality rates (5.6% vs. 17.4 % at 20 years; P < 0.001). Compared with the conventional treatment group, the early surgery group also had a significantly lower risk of overall mortality (HR, 0.66; 95% CI, 0.47 to 0.93; P = 0.018) and significantly lower overall mortality rates (28.2% vs. 33.9% at 20 years; P = 0.022).

**Conclusions:** Compared with conventional management, early surgery is associated with reduced long-term cardiac and overall mortality among asymptomatic patients with severe MR and preserved left ventricular function (ClinicalTrials.gov number, NCT01703806).

---

Surgery is recommended for severe, degenerative mitral regurgitation (MR) in symptomatic and asymptomatic patients with left ventricular (LV) dysfunction (1,2). However, controversy remains as to whether mitral valve (MV) repair should be performed in asymptomatic patients without LV dysfunction (3–5), and the current guidelines differ on indications for early surgery. The current American Heart Association/American College of Cardiology (AHA/ACC) guidelines recommend early surgery for asymptomatic patients with preserved LV function, if the likelihood of a successful MV repair is >95%, but the 2021 European Society of Cardiology (ESC) guidelines recommend watchful waiting for such patients (1,2).

The clinical outcomes of asymptomatic severe MR are heterogeneous (6), and asymptomatic patients with a flail MV or a larger effective regurgitant orifice area (EROA) of MR have shown poor outcomes under medical management (7,8). Among asymptomatic patients with flail MV, early surgery, compared with initial medical management, was associated with a significant survival benefit (9). However, according to a small observational study (10), the overall survival of such patients managed with a watchful waiting strategy was comparable to the expected survival of the general population for up to 15 years. These controversial results underscore persistent uncertainty. Although a randomized clinical trial could provide definitive evidence, it is highly unlikely that a large, randomized trial will ever be conducted for asymptomatic MR. Data from large, carefully designed registries are needed to improve quality of care (1), and we previously showed that early surgery was associated with reduced cardiac mortality, but not overall mortality in asymptomatic severe MR (4,11). In the present study, we enrolled more than 1,000 asymptomatic patients with degenerative MR (EROA of MR > 0.4 cm^2^) and extended the duration of follow-up to > 20 years. We sought to examine the hypothesis that early surgery is associated with better survival in these patients by comparing long-term cardiac mortality and all-cause mortality between early surgery and conventional treatment with the use of propensity score adjustment.

## METHODS

### Study Design

We conducted this prospective, multicenter, observational study involving asymptomatic patients with severe degenerative MR who were candidates for either early surgery or conservative management at two centers in Korea. The study protocol was designed by the principal investigator and approved by the institutional review board of each participating center. The first draft of the manuscript was prepared by the first two authors and was reviewed and edited by all the authors. The first authors and corresponding author made the decision to submit the manuscript for publication and vouch for the fidelity of this report to the protocol and for the accuracy and completeness of the data and analyses.

### Patient Selection

Patients were eligible for enrollment if patients aged 20 to 85 years; had severe, degenerative MR without exertional dyspnea; had an LV ejection fraction (EF) greater than 60%; had an end-systolic dimension (ESD) smaller than 40 mm. Severe, degenerative MR should fulfill the following criteria: severe MV prolapse with loss of coaptation or flail leaflet and holosystolic MR whose EROA larger than 0.4 cm^2^ (1).

In accordance with the 2006 ACC/AHA guidelines on surgical indications for severe MR (12), patients were excluded if they had exertional dyspnea or angina, an LV EF ≤ 60%, an LV ESD ≥ 40 mm, new onset of atrial fibrillation secondary to MR, or significant aortic valve disease.

### Study Procedures

Eligibility was determined after each patient underwent a thorough evaluation of symptom status, medical records, and results of echocardiography. After enrollment in the study, the treatment groups were not assigned randomly. Instead, the risks and benefits of early surgery and conventional treatment were explained to each patient, and the patients and attending physician collaborated to choose a management option that aligned with the individual patient’s preferences and values, sharing responsibility for the decision of treatment. Informed consent was obtained from each patient and the study protocol was approved by the ethics committee of our institution.

The protocol specified that patients in the early surgery group should undergo MV surgery within 6 months of enrollment. Patients in the conventional treatment group were treated according to the AHA/ACC guidelines,(1,12) and were referred for surgery if they became symptomatic during follow-up, if the LV EF ≤ 60%, LV ESD ≥ 40 mm, or Doppler estimated pulmonary artery pressure > 50 mmHg on follow-up echocardiography, or if atrial fibrillation developed.

Echocardiographic evaluation was performed at baseline and annually during follow-up. End-diastolic dimension (EDD) and ESD of the LV were measured from parasternal M-mode acquisitions, and end-systolic volume, end-diastolic volume and EF of the LV were calculated with the biplane Simpson method (13). Comprehensive echocardiographic evaluation of MR was performed using the integrated approach of two dimensional, Doppler and color flow imaging. The EROA was determined by dividing the regurgitant flow rate calculated as 2πr^2^ × aliasing velocity, where r is the proximal isovelocity surface area (PISA) radius, by peak MR velocity (14). A regurgitant volume was estimated as EROA multiplied by the velocity time integral of the MR jet. In cases of eccentric or multiple MR jets for which PISA method is less accurate, regurgitant volume and EROA were also obtained by volumetric methods based on quantitative Doppler measurement of mitral and aortic stroke volume (15).

The protocol specified that all the study patients would be followed at 3 months, 6 months, and 1 year after enrollment, and at 6-month intervals thereafter until close-out of the study. They were also educated to report to a study coordinator or an investigator if they experienced exertional dyspnea or any adverse events. Adherence to scheduled follow-up was complete for 839 (79%) patients, whereas 224 (21%) patients refused to visit the heart valve clinic regularly, preferring to visit the clinic only when they experienced symptoms or events. For the 38 (7%) patients in the conventional treatment group and the 17 (3%) patients in the early surgery group who were lost to follow-up, data on vital status, dates and causes of death were obtained from the Korean national registry of vital statistics. The primary end point was cardiac death during follow-up, and the secondary end point was all-cause death during follow-up.

### Statistical Analysis

Analyses followed the intention-to-treat principle, where all study patients were included in the treatment group which they originally selected at the time of enrollment. Baseline clinical and echocardiographic characteristics were compared in the two treatment groups using Student’s *t* test or the Mann-Whitney U test for continuous variables and the chi-square test or Fisher’s exact test for categorical variables as appropriate. Event-free survival curves were constructed with Kaplan-Meier estimates and compared using the log-rank test. Cox proportional hazards analysis was performed to compare hazard rates of outcomes between the early surgery and conventional treatment group. Given the potential imbalance in baseline characteristics between enrolled patients in the treatment groups, outcomes were compared with the use of propensity score adjustment (propensity score matching and overlap propensity score weighting) to correct for selection biases and potential confounding (16,17). The propensity scores were estimated without regard to outcome variables, using multiple logistic-regression analysis. All prespecified covariates were included in the full non-parsimonious models for treatment with early surgery versus conventional strategy (Table 1). The discrimination and calibration ability of the propensity score model was assessed by means of the C statistic (C = 0.73) and the Hosmer-Lemeshow statistic (chi-square = 3.67, df =8, p = 0.89). The propensity score-matched pairs were created by matching early surgery and conventional strategy subjects on the logit of the propensity score using calipers of width equal to 0.2 of the standard deviation of the logit of the propensity score (18). After propensity-score matching, we examined the similarity of early surgery and conventional strategy subjects in the propensity score matched sample by calculating standardized differences for each of the baseline variables listed in Table 1. All of the standardized differences for each of the baseline variables were less than 0.10 (10%) after matching. In the propensity score-matched cohort, the risks of mortalities were compared using Cox regression models with robust standard errors that accounted for the clustering of matched pairs. Moreover, we also adjusted for differences in baseline characteristics in the overall cohort by using overlap propensity score weighting. Overlap weighting is a propensity score method, in which outliers do not dominate results and worsen precision (17). Additionally, the consistency of treatment effect was evaluated with tests for interaction among subgroups according to compliance with the study protocol. All reported p values were 2 sided, and a p value <0.05 was considered statistically significant. SAS software, version 9.1 (SAS Institute, Inc., Cary, North Carolina) and R package, version 3.3.1 were used for statistical analyses.

**Table 1.** Baseline Clinical and Echocardiographic Characteristics of the Study Patients According to Treatment Group.

| Characteristics | Overall cohort |  |  |  | Propensity Score-Matched Cohort |  |  |
| --- | --- | --- | --- | --- | --- | --- | --- |
|  | CONV<br>(N=518) | Early Surgery<br>(N=545) | p-value<sup>1</sup> | Standardized<br>Difference of Mean (%) | CONV<br>(N=358) | Early Surgery<br>(N=358) | Standardized<br>Difference of Mean(%) |
| Age, years | 50.5 ± 14.2 | 52.2 ± 13.1 | 0.038 | -12.7 % | 51.6 ± 14.5 | 51.2 ± 13.7 | -4.20 % |
| Male gender, n (%) | 340 (65.6) | 333 (61.1) | 0.125 | 9.4 % | 221 (61.7) | 224 (62.5) | -1.73 % |
| Body surface area (m<sup>2</sup>) | 1.73 ± 0.18 | 1.73 ± 0.19 | 0.991 | -0.1 % | 1.72 ± 0.18 | 1.72 ± 0.19 | 0.53 % |
| Body mass index (kg/m<sup>2</sup>) | 23.9 ± 3.3 | 24.4 ± 3.4 | 0.017 | -14.7 % | 24.0 ± 3.2 | 24.1 ± 3.3 | -2.37 % |
| Smoking, n (%) | 127 (24.7) | 136 (25.0) | 0.926 | -0.6 % | 89 (24.9) | 95 (26.5) | -3.84 % |
| Diabetic mellitus, n (%) | 32 (6.2) | 34 (6.2) | 0.967 | -0.3 % | 24 (6.7) | 18 (5.0) | 7.14 % |
| Chronic renal insufficiency, n (%) | 2 (0.4) | 2 (0.4) | >0.999 | 0.3 % | 1 (0.3) | 1 (0.3) | 0.00 % |
| Hypertension, n (%) | 185 (35.7) | 200 (36.7) | 0.739 | -2.1 % | 115 (32.1) | 118 (33.0) | -1.79 % |
| Atrial fibrillation, n (%) | 4 (0.8) | 14 (2.6) | 0.023 | -14.1 % | 4 (1.1) | 4 (1.1) | 0.00 % |
| Drug therapy, n (%) |  |  |  |  |  |  |  |
| Angiotensin receptor blocker | 88 (17.0) | 162 (29.7) | <0.001 | -30.5 % | 70 (19.6) | 73 (20.4) | -2.10 % |
| ACE inhibitor | 30 (5.8) | 37 (6.8) | 0.499 | -4.2 % | 21 (5.9) | 19 (5.3) | 2.43 % |
| Calcium channel blocker | 48 (9.3) | 71 (13.0) | 0.052 | -12.0 % | 33 (9.2) | 30 (8.4) | 2.96 % |
| Beta-blocker | 43 (8.3) | 85 (15.6) | <0.001 | -22.6 % | 38 (10.6) | 30 (8.4) | 7.63 % |
| Diuretics | 61 (11.8) | 135 (24.8) | <0.001 | -34.1 % | 55 (15.4) | 57 (15.9) | -1.54 % |
| Nitrate | 15 (2.9) | 16 (2.9) | 0.969 | -0.2 % | 12 (3.4) | 8 (2.2) | 6.78 % |
| Statin | 28 (5.4) | 60 (11.0) | <0.001 | -20.5 % | 24 (6.7) | 22 (6.2) | 2.28 % |
| EuroSCORE | 1.06 ± 1.39 | 1.20 ± 1.41 | 0.041 | -10.3 % | 1.13 ± 1.45 | 1.12 ± 1.38 | 0.99 % |
| Charlson comorbidity index | 0.71 ± 1.21 | 0.96 ± 1.10 | <0.001 | -21.7 % | 0.79 ± 1.31 | 0.81 ± 1.01 | -1.43 % |
| Reasons for echocardiography |  |  |  | 44.9 % |  |  | 6.24 % |
| Cardiac murmur alone | 74 (14.3) | 54 (9.9) |  |  | 41 (11.5) | 46 (12.9) |  |
| Referral from other hospital | 310 (59.9) | 430 (78.9) |  |  | 260 (72.6) | 255 (71.2) |  |
| Pre-operative consultation* | 16 (3.1) | 11 (2.0) |  |  | 11 (3.1) | 11 (3.1) |  |
| Cardiovascular health checkup | 118 (22.8) | 50 (9.2) |  |  | 46 (12.9) | 46 (12.9) |  |
| Echocardiographic characteristics |  |  |  |  |  |  |  |
| End-systolic dimension, mm | 34.2 ± 3.8 | 35.1 ± 4.3 | 0.001 | -20.0 % | 34.7 ± 3.7 | 34.7 ± 4.3 | 0.33 % |
| End-diastolic dimension, mm | 57.1 ± 5.2 | 58.9 ± 5.5 | <0.001 | -34.0 % | 57.9 ± 5.0 | 58.1 ± 5.2 | -3.97 % |
| Left atrial dimension, mm | 45.3 ± 6.9 | 47.0 ± 6.4 | <0.001 | -24.7 % | 46.0 ± 7.2 | 46.1 ± 6.5 | -1.14 % |
| Left ventricular ejection fraction, % | 66.0 ± 4.7 | 66.4 ± 4.9 | 0.125 | -9.4 % | 65.8 ± 4.7 | 66.1 ± 4.9 | -5.81 % |
| EROA of MR (cm<sup>2</sup>) | 0.73 ± 0.31 | 0.79 ± 0.31 | <0.001 | -19.3% | 0.76 ± 0.33 | 0.76 ± 0.29 | 0.64 % |
| Prolapsed segment |  |  | 0.178 | 10.4 % |  |  | 0.00% |
| Anterior | 111 (21.4) | 97 (17.8) |  |  | 73 (20.4) | 71 (19.8) |  |
| Posterior | 280 (54.1) | 324 (59.5) |  |  | 199 (55.6) | 201 (56.2) |  |
| Both | 127 (24.5) | 124 (22.8) |  |  | 86 (24.0) | 86 (24.0) |  |

Values are mean ± SD or n (%)
\* Preoperative consultation for noncardiac surgery
ACE denotes angiotensin-converting enzyme; CONV, Conventional Treatment Group; EROA, effective regurgitant orifice area; EuroSCORE, European System for Cardiac Operative Risk Evaluation score; MR = Mitral regurgitation

## RESULTS

### Patient Characteristics

From 1996 to 2016, a total of 1,063 consecutive asymptomatic patients with severe degenerative MR and preserved LV function were enrolled. The mean (±SD) age of the patients was 51.4±13.7 years and 63.3% were men. Early surgery was performed on 545 patients and the conventional treatment strategy was chosen for 518 patients. The baseline clinical and echocardiographic characteristics of the early surgery group and the conventional treatment group were compared, as shown in Table 1. There were no significant differences between the two groups in terms of gender, body surface area, smoking, diabetes mellitus, hypertension, and EF. However, the early surgery group was older, had a higher comorbidity index and larger EROA of MR, LV ESD and EDD, and left atrial diameter (P < 0.01). Propensity-score matching for the overall cohort yielded 358 matched pairs of patients. In the matched cohort, there were no longer any significant differences between the early surgery and conventional treatment groups for any covariates, according to the use of statistical methods appropriate for matched data (Table 1).

### Surgical Procedures

The procedures were performed with the use of standard cardiopulmonary bypass. In the early surgery group, all the patients underwent surgery within 6 months after enrollment; the median time between enrollment and surgery was 29 days (interquartile range, 8.0 to 53.8). MV repair or replacement was performed successfully in 528 (96.9%) and 17 (3.1%) patients, respectively, and there were no cases of operative mortality in the early surgery group. Concomitant maze operation or tricuspid valve repair was performed in 19 patients (3.5%) and 33 (6.1%), respectively, while coronary artery bypass grafting (CABG) operation was carried out in 33 (6.1%). In the conventional treatment group, 234 patients underwent late MV surgery with MV repair (n = 197) or MV replacement (n = 37), and 3 patients were treated with transcatheter edge-to-edge repair of MV (n = 3) at a mean interval of 6.4 ± 4.8 years after enrollment during follow-up. Concomitant maze operation, tricuspid valve surgery and CABG operation were performed in 59 patients (25.2%), 34 (14.5%) and 13 (5.6%), respectively. There were two (0.8%) operative mortalities in the conventional treatment group with late surgery, and the repair rate was significantly lower than that in the early surgery group (84.2% vs. 96.9%, P < 0.01).

### Comparison of Outcomes in the Overall Cohort

Data collection ended in October 2024. The median follow-up was 12.0 years (interquartile range, 8.3 to 16.8). In an intention-to-treat analysis including all the study patients, 8 (1.5%) of 545 patients who underwent early surgery and 54 (10.4%) of 518 patients who chose conventional management died from cardiovascular causes (hazard ratio [HR] in the early-surgery group, 0.17; 95% confidence interval [CI], 0.07 to 0.40; P < 0.001) (Table 2). The cumulative incidence of the primary end point (cardiac mortality), as adjusted with the use of overlap weighting, was 0.4 ± 0.2% at 10 years and 5.6 ± 2.3% at 20 years in the early surgery group as compared with 5.5 ± 1.0% at 10 years and 17.4 ± 2.3% at 20 years in the conventional management group (P < 0.001) (Fig. 1A). A total of 74 deaths (13.6%) from any cause occurred in the early surgery group and 116 (22.4%) in the conventional management group (HR, 0.72; 95% CI, 0.52 to 0.99; P = 0.046). The estimated actuarial mortality rates were significantly lower in the early surgery group than in the conventional management group (4.5% vs. 11.3% at 10 years and 29.6% vs. 33.9% at 20 years; P = 0.034) (Fig. 1B). In the secondary per-protocol analysis that included 839 patients who completed scheduled follow-up, the early surgery group had a lower risk of cardiac mortality (HR, 0.16; 95% CI; 0.05 to 0.53) and all-cause mortality (HR, 0.83; 95% CI; 0.56 to 1.25) than the conventional treatment group. Among the 224 patients with incomplete adherence to scheduled follow-up, the early surgery group also had a lower risk of cardiac mortality (HR, 0.21; 95% CI; 0.07 to 0.61) and all-cause mortality (HR, 0.51; 95% CI; 0.30 to 0.88). The subgroup analysis according to adherence to scheduled follow-up found no significant interactions in risk of cardiac mortality (P for interaction = 0.73) and all-cause mortality (P for interaction = 0.16) between the treatment groups, suggesting that the survival benefits of early surgery were consistent regardless of compliance with the study protocol.

**Figure 1.**
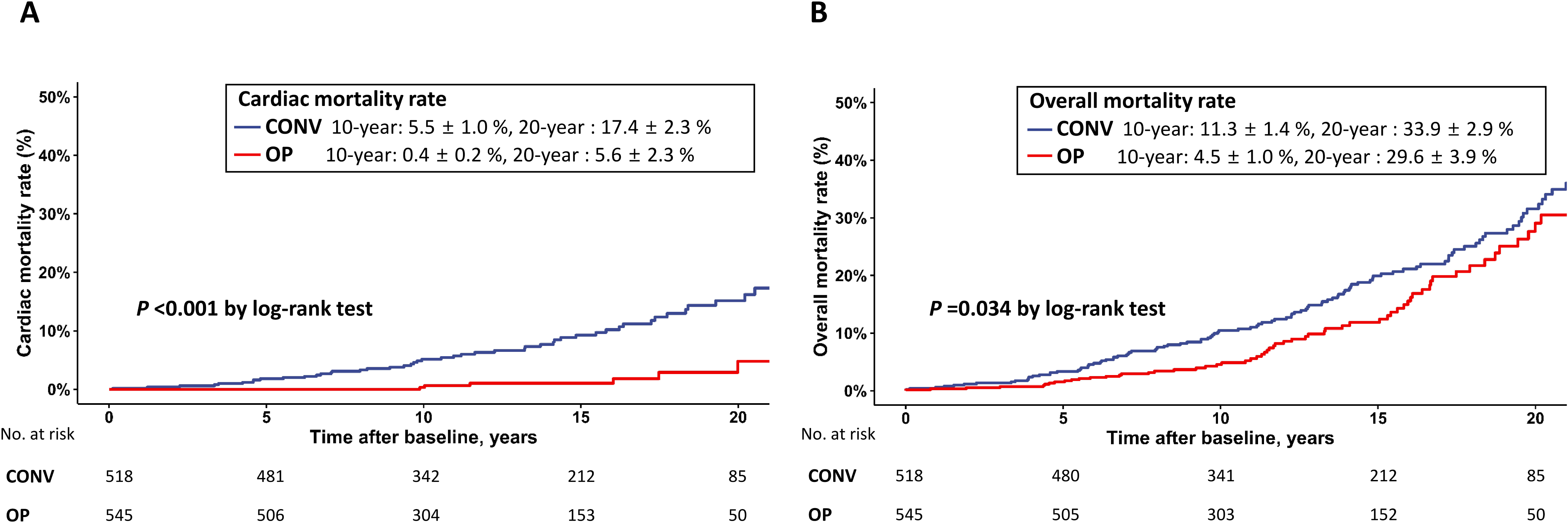
Comparison of conventional treatment (CONV) and early surgery (OP) in terms of actuarially determined cardiac mortality rates (A) and overall mortality rates (B), adjusted by overlap weighting, in the overall cohort

**Table 2.** Cox Proportional Hazards Regression Analysis for Mortality in Early Surgery Versus Conservative Treatment in the Overall Cohort.

|  | Number of Events |  | Adjusted by Overlap Weighting |  |
| --- | --- | --- | --- | --- |
|  | Early Surgery<br>(n=545) | Conventional Treatment<br>Group<br>(n=518) | HR (95% CI) | P-value |
| All-cause Death | 74 | 116 | 0.716 (0.515 to 0.994) | 0.046 |
| Cardiac Death | 8 | 54 | 0.172 (0.074 to 0.402) | <0.001 |

In the early surgery group, 38 (7.0%) patients had recurrences of moderate (n = 30) or severe MR (n = 8), and 8 (1.5%) underwent repeat MV surgeries. The estimated actuarial 20-year rates of MR recurrence and repeat MV surgery in the early surgery group were 8.5 ± 1.4% and 3.7 ± 1.5%, respectively. Atrial fibrillation developed in 73 patients (13.4%) during follow-up.

### Comparison of Outcomes in the Propensity-matched Cohort

For the 358 propensity-score matched pairs, the median follow-up was 12.0 years (interquartile range, 8.1 to 16.7) in the early-surgery group and 13.3 years (interquartile range, 9.0 to 17.5) in the conservative-management group (P = 0.13). During follow-up, 6 patients (1.6%) in the early surgery group and 37 (10.3%) in the conventional management group died from cardiac causes (Table 3). The early surgery group had a significantly lower risk of cardiac mortality than the conventional treatment group (HR 0.18; 95% CI, 0.08 to 0.43; P < 0.001) and a significantly lower 20-year cardiac mortality rate (5.6 ± 2.8% vs. 17.4 ± 3.1%, P < 0.001; Figure 2A). A total of 48 deaths (13.4%) from any cause occurred in the early-surgery group and 80 (22.3%) in the conventional-management group. Compared with the conventional treatment group, the early surgery group had a significantly lower risk of overall mortality (HR 0.66; 95% CI, 0.47 to 0.93; P = 0.018), and the estimated actuarial mortality rates were significantly lower in the early-surgery group than in the conservative-management group (3.9% vs. 11.2% at 10 years and 28.2% vs. 33.9% at 20 years; P = 0.022) (Figure 2B). In the subgroup analysis according to adherence to scheduled follow-up, there were no significant interactions between the treatment group and the risk of cardiac mortality (P for interaction = 0.68) and all-cause mortality (P for interaction = 0.36).

**Figure 2.**
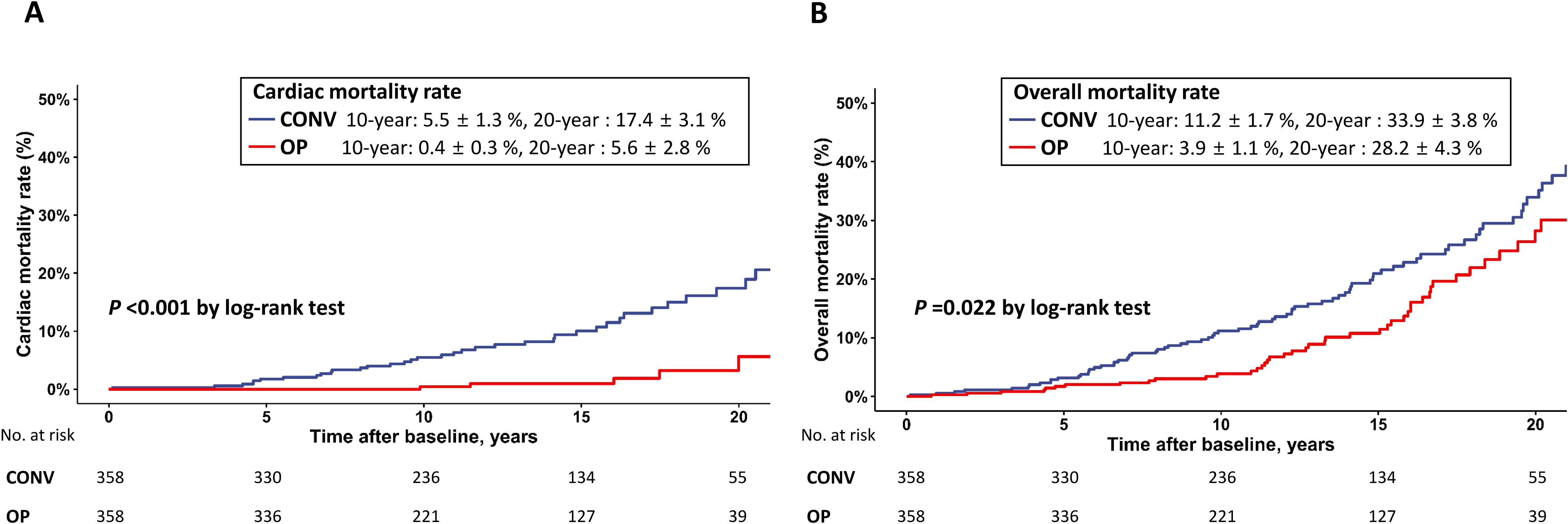
Comparison of conventional treatment (CONV) and early surgery (OP) in terms of actuarially determined cardiac mortality rates (A) and overall mortality rates (B) in the propensity-matched cohort

**Table 3.** Cox Proportional Hazards Regression Analysis for Mortality in Early Surgery Versus Conservative Treatment Group in the Propensity Score-Matched Cohort.

|  | Number of Events |  | Propensity Score Matching |  |
| --- | --- | --- | --- | --- |
|  | Early Surgery<br>(n = 358) | Conventional Treatment<br>Group<br>(n=358) | HR (95% CI) | P-value |
| All-cause death | 48 | 80 | 0.660 (0.468 to 0.930) | 0.018 |
| Cardiac death | 6 | 37 | 0.179 (0.075 to 0.428) | <0.001 |

## DISCUSSION

The present study demonstrates that compared with conventional treatment, early surgery is associated with significant long-term reductions in overall mortality and cardiac mortality among asymptomatic patients with severe MR and preserved LV systolic function. In our previous studies, we examined the hypothesis that early surgery is associated with a significant decrease in all-cause and cardiac mortality but did not find a significant association between early surgery and improved survival (4,11). Enrollment of > 1,000 study patients and extending follow-up duration to > 20 years provided our study with sufficient power to detect a significant difference in overall mortality between the two management strategies in a propensity analysis. Analysis of the Mitral Regurgitation International Database (MIDA) registry also showed that long-term survival rates were significantly higher in patients with early surgery than those with initial medical management (86% vs 69% at 10 years), which was confirmed in a propensity-matched cohort (9). In contrast, watchful observation with a protocol of clinical and echocardiographic follow-up in the heart valve clinic may have improved outcomes of initial medical management (3), and a small observational study reported an overall survival rate of 85.6% at 10 years for patients managed with careful surveillance (10), which was similar to the survival rate of those who underwent early surgery in the MIDA registry. Therefore, there is no consensus on the optimal management strategy (5). In the present study, overall survival in the conventional treatment group, which also followed a surveillance protocol, was 89% at 10 years, suggesting a similarly favorable prognosis of asymptomatic MR with careful surveillance. Our findings might appear to support the recommendation of the ESC guidelines that MV surgery can be safely deferred until symptoms or LV dysfunction (LV ESD > 40 mm or LV EF <60%) develop in asymptomatic patients with severe MR (2). However, our results also present a compelling case for early surgery, potentially challenging the conventional watchful observation approach, because the early surgery group had higher overall survival rates (96% at 10 years) than the conventional treatment group, as confirmed in a propensity-matched cohort. To our knowledge, our study provides the strongest evidence supporting the current AHA/ACC guidelines, which recommend early surgery for asymptomatic patients with preserved LV function and EROA of MR > 0.4 cm^2^ (1), because a randomized clinical trial involving more than 1,000 trial patients and requiring a follow-up duration of > 20 years would be infeasible.

Several prerequisites should be met before considering early surgery in asymptomatic patients with severe MR. First, early surgery should be performed at valve centers with > 95% likelihood of successful MV repair and with an expected operative mortality of < 1% (1); in our early surgery group, the repair rate was 97%, and operative mortality was 0%. A recent cohort study using The Society of Thoracic Surgeons Adult Cardiac Surgical Database also reported that operative mortality for MV repair and risk of conversion to MV replacement were lower at higher-volume centers with an operative mortality of 0.73% and conversion rate of 3.69% (19). In contrast, the repair rate was 84% in the conventional treatment group of the present study, and an outcome study of the watchful waiting strategy reported that MV repair was performed in 86% of patients, with a reoperation rate of 6.3% due to failed repairs (10). Second, a durable repair should be provided with 95% freedom from reoperation and >80% freedom from recurrence of moderate or severe MR at 15 to 20 years after surgery (1,20,21); the rates of MR recurrence and repeat MV surgery at 20 years were 8.5 ± 1.4% and 3.7 ± 1.5%, respectively, in the early surgery group of the present study. In a recent report from Toronto General Hospital involving 1,234 patients who had MV repair, the probability of recurrent moderate or severe MR was 12.5% at 20 years and the cumulative incidence of reoperation on the MV was 4.6% at 20 years (22). Interestingly, the probability of developing new atrial fibrillation was 32.4% at 20 years, and atrial fibrillation was associated with cardiac and non-cardiac death in that study (22). As progressive remodeling of the left atrium associated with severe MR fosters the development of atrial fibrillation, early correction of MR may decrease the risk associated with new-onset atrial fibrillation. Third, quantitative assessment of MR has an important prognostic value (23, 24), and the EROA of MR should be greater than 0.4 cm^2^. Enriquez-Sarano et al. reported that asymptomatic patients with EROA of MR > 0.4 cm^2^ had excess mortality and that a survival benefit was associated with surgery for larger EROAs (8). The present study is the first to demonstrate a greater survival benefit of early surgery in asymptomatic patients with EROA > 0.4 cm^2^ by directly comparing early surgery with watchful waiting strategy. It should be pointed out that enrollment criterion for severe, degenerative MR in the present study was severe MV prolapse with loss of coaptation or flail leaflet, with EROA of holosystolic MR > 0.4 cm^2^. Fourth, a strategy of watchful waiting involves clinical and echocardiographic follow-up at a heart valve center every 6 months (10), and patient compliance with surveillance should also be considered for the decision of treatment. Considering that early MV repair eliminates the need for surveillance and also obviates the possibility of loss to follow-up (1,25), early surgery could be the preferred strategy for patients who are non-compliant with surveillance. In the present study, compared with conventional treatment, the subgroup of patients who did not adhere to the study protocol experienced a greater reduction in the risk of all-cause mortality than the compliant subgroup, implying that the advantage of early surgery would be greater in patients without compliance. Considering that a community cohort study has revealed substantial undertreatment of severe MR (26), early surgery might provide a much greater benefit than watchful waiting strategy in real-world practice.

### Study Limitations

The present study was subject to limitations resulting from the nonrandom assignment of treatment groups, which may have influenced the results due to selection bias and potential confounding. Propensity score matching and overlap weighting attempt to mimic important attributes of randomized clinical trials and adjust for biases related to treatment selection and imbalance in baseline characteristics (16,17). Although residual confounding cannot be completely excluded, our robust study design and propensity-based statistical analyses strengthen the internal validity of our results.

Incomplete adherence to scheduled follow-up might have delayed recognition of surgical triggers in the conventional treatment group, although the subgroup analyses according to compliance found no significant interactions in outcomes between the treatment groups. Because operative mortality was very low and the MV repair rate was 97% in the early surgery group, our results may not be applicable to low-volume medical centers or patients at high operative risk.

## Conclusion

Compared with conventional management, early surgical correction of MR is associated with reduced long-term cardiac and all-cause mortality among asymptomatic patients with severe MR. The survival benefit of early surgery was confirmed in a propensity-matched cohort. Although conventional management appears to be a safe strategy, early surgery may be a more effective treatment option for asymptomatic patients with severe MR at excellent heart valve centers.

## Data Availability

Because of data privacy restrictions, data from the present study are unable to be shared outside of the aurhors of the present study.

